# Improving US Virgin Islands’ heath system: “You get more benefits with your insurance in the States than you get here”

**DOI:** 10.1101/2020.07.02.20135657

**Authors:** Karen H. Wang, Zoé M. Hendrickson, Hannah R. Friedman, Maxine A. Nunez, Marcella Nunez-Smith

## Abstract

**Background:** The US Virgin Islands (USVI) are actively rebuilding their healthcare delivery system following destruction by Hurricanes Irma and Maria in 2017.

**Methods:** In 2013, we conducted a qualitative study in the US Virgin Islands using semi-structured one-on-one interviews to explore individuals’ decision-making regarding healthcare-seeking off-island. The coding team analyzed the transcripts using a constant comparative analysis, and Atlas.ti to organize our emerging thematic analysis.

**Results:** Five themes emerged from 19 interviews that illustrate healthcare system level factors that influence participants’ decisions about seeking healthcare off-island: 1) limited availability of services and desire for options, 2) limited accessibility of services, 3) healthcare system interactions and experiences, 4) healthcare system policies, and 5) trust in healthcare systems.

**Conclusions:** The experiences of care seeking off-island for our sample highlight several mechanisms through which the USVI healthcare delivery system could improve, including the adoption of telemedicine, changes in insurance, and healthcare workforce policies.

---

In 2017, Hurricanes Irma and Maria destroyed the healthcare infrastructure of islands throughout the Caribbean region, including the United States Virgin Islands (USVI), an archipelago located in the Eastern Caribbean. The USVI consists of three main islands––Saint Croix, Saint Thomas, and Saint John––and has been a territory of United States since 1917. Physical damage to the already vulnerable healthcare delivery systems severely limited access to and utilization of services. A six-month post hurricane report examined federally funded health centers in the USVI and Puerto Rico to assess their current needs. The report found that many were operating at partial capacity, in physical disrepair, and with significant human resource challenges, both in hiring and retention.^1^ Primary challenges reported by the health centers included staff loss of homes, lack of reliable transportation and usable roads, and staff members’ plans to leave Puerto Rico or USVI. A year after the hurricanes, USVI’s two main hospitals needed complete reconstruction, while some of Puerto Rico’s hospitals and community health centers were operating on generators or limited grid power.^1-3^ This destruction of infrastructure has impacted health access and outcomes. One recent study documented an increase in emergency rooms visits from diabetes and asthma in the year after the hurricanes.^4^ Worsening outcomes for individuals with chronic diseases after the hurricanes has been partly attributed to unstable facilities rendering poor access to care and supply of medications.^5^

The islands throughout the region continue substantive efforts to rebuild and strengthen their health systems. As these efforts continue, there is an opportunity to draw on existing knowledge about the health needs and preferences of the region’s residents to improve the healthcare systems moving forward. A handful of studies conducted prior to the 2017 hurricanes in the Caribbean have shown that individuals living in the Caribbean region had high rates of chronic diseases such as cardiovascular disease, cancer, diabetes, and lung disease.^6-8^ Despite the increased disease burden in the Caribbean, there is a limited body of research on healthcare quality and outcomes focused on the US territories of USVI and Puerto Rico. Two studies demonstrated sub-optimal quality of care in the territories, including higher hospital readmission rates for pneumonia and acute myocardial infarction and documented delays in appropriate breast cancer care.^9-11^

There is limited research on perspectives of individuals in the Caribbean territories about their concerns and experiences with the healthcare system. One qualitative study exploring the main health issues facing USVI residents found that participants were highly concerned about rising burden of chronic diseases, the high costs of on island-care, the limited resources available to improve healthcare USVI, and the quality and training of healthcare providers.^12^ With the exception of this prior research study, there are no other studies focused on community members’ experiences and perspectives about the USVI healthcare system. Building on this prior research, we explored the experiences and preferences of residents of the USVI seeking healthcare in different locations, specifically those traveling off-island to obtain healthcare. This data can provide unique insights to inform the rebuilding efforts in this region and strengthen the healthcare delivery system.

## Methods

### Study Design, Setting, and Sample

We conducted a qualitative study to explore the experiences and decision-making processes of individuals accessing healthcare by crossing territorial, regional, or national borders. We emailed a recruitment letter to individuals with a University of Virgin Islands email address and recruited a convenience sample of individuals forty years or older living in the USVI who had accessed healthcare services across regional or national borders.

Our study team consisted of method and content experts on healthcare access and utilization in the USVI. We developed an interview guide that asked participants about their motivations for accessing healthcare in other locations, the challenges they faced, and their perspectives on this type of movement and its effect on their health.

One member of the study team (XX) trained in qualitative methods conducted interviews in a private office on the campus of the University of the Virgin Islands, St Thomas, UVSI. We audio-recorded and professionally transcribed each interview and reviewed the recordings to ensure proper transcription. Interviews continued until we reached thematic saturation about participants’ experiences and perspectives. Interviews were conducted in March and May of 2013. Interviews lasted approximately 45 minutes to one and half hours. The Institution Review Board of Yale University approved this study.

### Data Analysis

The team developed a code structure in accordance with principles of grounded theory, using systematic, inductive procedures, and generated insights grounded in the views expressed by participants.^13,14^ Three members of the research team (XX, XX, XX) created the initial code structure by independently coding the transcripts, then meeting to negotiate consensus over differences in independent coding. The three researchers used the constant comparative method to ensure that emerging themes were consistently classified, to refine and expand on existing codes, and to identify novel concepts.^13^ Through this process, we achieved consensus and finalized a comprehensive code structure capturing all data concepts (XX, XX, and XX) refined the code structure with feedback from (XX and XXX) and systematically applied the final code structure to all transcripts. We organized the codes into the individual-, interpersonal-, organizational level factors influencing decisions on where to obtain healthcare services. We wrote memos on each code to identify patterns from the data, explore relationships between codes, and summarize our analysis of the data for each code. We then had multiple conversations around each memo to clarify and interpret our findings. We used qualitative analysis software (ATLAS.ti 5.0, Scientific Software Development, Berlin, Germany) to facilitate data organization and retrieval.

## Results

We highlight five emerging themes from the nineteen interviewees that characterized the organizational or healthcare system-level factors influencing people’s decisions about where they go for healthcare: 1) limited availability of services and desire for options, 2) limited accessibility of services, 3) healthcare system interactions and experiences, 4) healthcare system policies, and 5) trust in healthcare systems.

### Theme 1

*Participants wanted options from which to choose a healthcare provider, with a lack or limited availability of services on-island influencing their decisions to seek care off-island*. Many participants described that on-island healthcare options were limited compared to options off-island, and that these limitations specifically related to specialty healthcare personnel and services. On-island healthcare limits the number of providers a patient can select from, whereas off-island healthcare enables participants to choose from more than one healthcare provider. Participants often used healthcare services off-island because of this increased availability of multiple providers.

USVI’s geography and population size were considered by many as a restrictive force in availability of services. These immutable aspects of the island limit the types of services that may be sustained on-island.

> *Being on an island maybe, …there’s only so much of an amount of machines you can bring in. We’re on an island. It’s not like you can drive from a state. You can’t drive from one place to the next to get other machines here. We have to wait for things to come in via the barge or catch a plane I guess*.
>
> *Woman, 51 years old*

Other participants also perceived a relationship between geographic isolation and substandard care, suggesting that as a general rule, the quality of care is worse in isolated areas than in large metropolitan areas. Participants drew similarities between USVI and rural areas or small towns of mainland US in terms of the suboptimal quality of care. As one participant stated,

> *We’ve been here enough years to know that the health care here is not up to snuff. I know it’s as good as in some rural areas in the US mainland*.
>
> *Man, 60 years old*

The lack of specialty care on-island was compared to rural mainland US.

> *We are in a small, isolated spot. I mean, it’s the same thing if I looked in a town of 60,000 people in Iowa, I think I would feel the same way*…*It’s just why [would] I go to an old – not necessarily old but a country doctor for something that you can go to a big hospital and have a specialist do*.
>
> *Woman, 51 years old*

Another participant recounted widespread usage of off-island services due to unavailability of specialty care services on-land and concerns about the quality of the care that is available. The participant also highlighted the ability to choose between providers on the mainland.

> *I mean there’s a lot – I think maybe 50% of the people going off island go because they can’t get something. You can’t get that type of care here. Like a lot of people will go off for laser eye surgery or go to see an eye specialist at or stuff like that…So, stuff like that. That people go a lot of times [they] go off to the States to see a specialist more than anything. I think we’re the only ones who go off island to get the physical. I think mostly its people going to see a specialist, but a lot of people do. Especially I heard like – I mean the cancer center here. I mean I know people who are closely associated with the cancer center who say, “Don’t ever try to go to them for help*.*” Other people who said, “They’re great. They’re wonderful*.*” So personally, I’d feel more comfortable going to the States. If dealing with something that serious I just feel more comfortable going to the States because if you don’t like them, you can go down the street to plan B*.
>
> *Woman, 41 years old*

Another participant attributed prioritization of geographic distance and type of healthcare service needed to their decision to obtain healthcare on- or off-island. In this case, the participant obtains healthcare in both on and off-island locations, preferring obtaining healthcare on-island if it is available and obtaining healthcare off-island if they need a ‘second opinion’. As the availability of services on-island improved over time, the participant chose to receive more care on-island.

> *Because we didn’t have a cardiologist here. They didn’t have one here, so I was flying to Washington DC for a couple of years before there was a cardiologist here. That’s when I finally stopped traveling that distance. Remember it has some specialization doctors and in other areas it’s just general so you have to fly out if you need special care or a doctor that specializes in an area. It’s getting better now though, it is. It’s a lot better based on then and now. because now if I have to go to the emergency room and there’s something’s wrong, I know there’s a cardiologist that will come into the emergency room and see me because there’s one on island. But back then, it would be that they have to fly you out. You have to fly out…They [healthcare facility off-island] were good but I felt that “No, I can get that same service closer to home” and so I went to Puerto Rico. Other than that, I just go to the cardiologist and internal doctor here. I’m basically back and forth depending if I feel like based on what they tell me. I need a second opinion at that time, then I might go to Puerto Rico or I might go to Florida. It all depends on how my health is on a regular basis*
>
> *Woman, 50 years old*

### Theme 2

*Participants’ decisions about healthcare are influenced by their ability to access services on- and off-island*. Access, which describes people’s ability to use healthcare services when and where they are needed, is affected by dimensions such as cost, distance, time, and ease of traveling to reach services ^15,16^.

For participants who were considering obtaining healthcare off-island, cost was a major factor that limited or facilitated individuals’ access to care. Healthcare on-island was considered expensive, while healthcare services off-island had charges associated with travel, including plane flights and hotels, that increased the cost.

> *From the medical side, I think it might be even cheaper than here. The expense comes in air travel and hotel stays. That’s where the expense is. As far as the actual medical care, it seems to be cheaper than it is here*.
>
> *Man, 50 years old*

Insurance policies, including co-pay and out-of-network costs, directly affect patient out-of-pocket costs and play a role in how participants make decisions. One participant explained how their insurance policy has changed over time as it pertains to on-vs. off-island healthcare coverage.

> *We’re part of that group insurance. I do remember there was a short period in there where they covered 80% of the procedure if it was on-island and only 60% if it was off-island. Of course, they don’t cover your travel. That was only for a few years and it certainly has not been the case for a long time. It’s just either you’re in the [insurance company] system or you’re not. Now, it’s 80 versus 60 if you go to a doctor that’s not a [insurance company] doctor. It doesn’t matter where you do it, that’s the same. No, I haven’t had any problems with that. Again they, of course, don’t cover your travel to go to the mainland*.
>
> *Man, 60 years old*

Participants talked about how insurance policies not only affect their out of pocket costs, but also the types of benefits and services they are able to obtain in different locations. One participant emphasized that on-island benefits are less than off-island.

> *You get more in the States. because this is a territory and the way how those programs are administered here is disproportionate to how they’re administered in the States. So that’s another motivational factor. You get more benefits with your [Insurance] in the States than you get here*.
>
> *Woman, 51 years old*

While some perceived healthcare to be cheaper off-island, often due to insurance coverage, some participants found extra costs that were not covered by insurance to be prohibitive to obtaining off-island healthcare.

> *Going out to see a doctor? I couldn’t financially afford it … You have to be able to afford it and at that time, no. I was fairly young. Did I have insurance? Yes, I had insurance, but insurance doesn’t pay for you to book passage, hotel, to go to a doctor away. It’s going to pay to cover your hospital, your doctor visit but it’s not going to pay for all the other costs that you need to pay if you’re going to do that. You try to get medical health at home*.
>
> *Woman, 50 years old*

Geographic proximity was also considered a barrier to accessing healthcare services. The distance necessary to travel to services available off-island meant that some participants did not seek care routinely off-island, or opted for travel to a more proximate location, such as Puerto Rico, rather than services in the mainland United States. One participant reflected on their experience receiving services in the mainland,

> *They were good, but I felt that “No, I can get that same service closer to home” and so I went to Puerto Rico. Other than that, I just go to the cardiologist and Internal doctor here*
>
> *Woman, 50 years old*

Finally, participants preferred how the health services off-island were co-located in one geographic location. This type of co-location increased participant’s sense of access to services and ease of using services off-island as compared to accessing services on-island in separate locations spread out geographically.

> *No, on-island it’s not like that. Of course, you’re going to different doctors but they’re going to be in different locations…It’s much more convenient doing it this way [going off-island]*.
>
> Man, 50 years old

### Theme 3

*Participants’ interactions and experiences with the healthcare system during scheduling appointments, wait times, and patient-provider visits, influenced decisions about where to obtain healthcare*. Participants’ interactions with the healthcare system affected their experiences within that setting and influenced subsequent decisions about where to seek healthcare. These interactions begin when planning or scheduling an appointment, and include in-person interactions in the facility, as well as interactions with healthcare providers between visits. Some participants indicated that “customer service” is important to the whole healthcare experience, influencing people’s decisions to obtain healthcare off-island. For example, a participant said,

> *They treat you like they’re glad to see you. They’re happy that you’re there…Ask anybody here who has been to the [Off-Island Healthcare setting] and they’ll tell you it’s awesome – the customer service, the care that they give you. They’re very attentive…Here? It’s like you’re bothering some of them here*.
>
> *Woman, 49 years old*

When planning and scheduling an appointment, participants valued speed and efficiency. One participant described delays when scheduling appointments on-island as related to availability of services.

> *We don’t have the availability of a lot of the specialty stuff and a lot of the advanced tests. Sometimes, even if they’re available here, sometimes it takes so long to get an appointment - a limited amount where you may only have one or two places doing a particular function. Stateside, there are so many other places you can get it done so it’s more expedient*.
>
> Man, 50 years old

Wait times upon arrival at the health facility were also considered a major challenge on on-island. Speed and efficiency were also preferred during healthcare visits with providers and follow-up on laboratory test results. Many participants emphasized positive experiences with having a “one-stop” model for the healthcare system, as this type of delivery facilitated efficiency. As this same participant expressed,

> *Well, I have family members and friends that go there and I actually accompanied a family member there. I realized that it was like a one-stop where you can have everything done within a day or two and you can get your results quickly and so forth, I started doing it. I liked the service there and I continued. Here, was a bit more difficult…your appointments may be days apart, or weeks apart or what have you. I thought it was just better to have a one-stop situation where you can get everything done or if you have a referral, you can get them at just a real timely fashion*.
>
> Man, 50 years old

During healthcare visits, providers also played instrumental roles in an individual’s decision of where to seek healthcare. Previous experiences or relationships with a provider, either negative or positive, affected whether a participant was comfortable seeking care on- or off-island.

> *I haven’t done it [preventive exam] there [off-island] but people I’ve known have been there and people have done it and it was smooth sailing. I haven’t done it there. I did it here and I didn’t like the effects of it after. So I said next time I will go off-island to do it*
>
> *Woman, 52 years old*

Participants often highlighted how providers themselves facilitated travel off-island for healthcare by referring them to services off-island, even if services were able on-island. One participant described their provider encouraging them to seek services off-island,

> *Even with my mother after going to [US mainland city] getting initial assessments and everything, treatment, we come down for follow-up. At one point I had an appointment with the doctor and the doctor says, “You should go back to [Us mainland city]*.
>
> *Woman, 41 years old*

### Theme 4

*Healthcare system structural factors affected where participants decide to obtain healthcare and how they experience seeking care off-island*. Participants described several mechanisms by which healthcare system structural factors, such as the mobility of healthcare workforce and health information systems, facilitate decisions around healthcare use off-island. These healthcare system structural factors also influence participants’ experiences and perceptions of the care.

Participants described the mobility of healthcare workforce as influencing their decisions on where to obtain healthcare services. The mobility of the healthcare providers both negatively and positively influence how participants viewed and experienced the healthcare system. For example, one participant described how healthcare providers travel to USVI and provide services on-island for several consecutive days each month. This delivery practice enables USVI residents to stay on-island for certain healthcare services that not often available on island.

> *he would come once a month or so…*.*so if you ever had something like an emergency I guess, you can figure that out. It worked much better. You could come on Saturdays or Sundays. He had a busy practice on his weekend trips. Maybe he came twice a month, I’m not even sure*.
>
> *Man, 49 years old*

Another participant described that the mobility of the healthcare workforce negatively impacted their perceptions of healthcare quality.

> *Maybe a lot of the travelling nurses and people that come through the hospital maybe this is just a job to them and they’re not – they don’t feel any connection to this culture or to the hospital because they’re just kind of in and out in three months. So, they just bide their time here. I don’t know. I don’t know. We have a lot of travelling nurses; that scares me*.
>
> *Woman, 41 years old*

Participants discussed how healthcare system structures influence their experience and use of healthcare off-island. For example, participants illustrated the coordination needed across multiple healthcare system entities, from providers, laboratories, pharmacies, and radiologic services, to enable continuity of care between different healthcare system entities separated by large geographic distances. They reported both positive and negative experiences with coordinating care across these systems, such as coordinating time-dependent laboratory blood draws on-island and analysis of results off-island.

Many participants had positive experiences with web-based patient portals to access their health information from off-island healthcare systems. More streamlined coordination and communication enhanced their overall healthcare experiences. Participants experienced more convenient and less costly healthcare services off-island, such as not having to make another office visit to obtain laboratory results. One participant described the speed and efficiency of such a platform available at an off-island healthcare system, which facilitated easy follow-up, communications between visits, and viewing test results.

> *They send you the records. You can go online and see your results in some instances, within a couple of hours. Some of your tests you can see your results live. They have follow-up visits plan, they’ll write you or send you an e-mail 30 to 45 day in advance letting you know your appointments are coming up*.
>
> Man, 50 years old

### Theme 5

*Trust in the healthcare system influenced where participants decided to obtain healthcare*. This theme of trust was directly influenced by other health system factors characterized by theme 1 through themes 4, such as the availability of healthcare services, experiences with and interactions with the healthcare system, and the perceived quality of healthcare infrastructure. Though participants used different terms, such as confidence, faith, or comfort, to articulate this sense of “trust,” all influenced their decisions on where/where not to obtain healthcare.

One participant described how the lack of availability of personnel on-island creates an environment of uncertainty across all sectors of work, including a lack of trust in the healthcare system.

> *Generally, it’s one person doing this. If that person goes sick, if that person is not in the office, then you have to wait because there’s only one person, one, one, one. You get that, not necessarily with healthcare, but with so many things within the community that I don’t trust the system; certainly, not for my healthcare*.
>
> *Woman, 62 years old*

Participants also described that their trust in the healthcare system comes from whether the infrastructure, equipment, or technology available appears state-of-the-art, and that their experiences influence how they perceive the healthcare system. One participant described that the lack of confidence in the USVI healthcare system may have roots in the perception that the overall infrastructure of USVI is “not great.”

> *Perhaps because so many of our social systems and our infrastructure here is not great, so you don’t have a lot of confidence in the medical system either*.
>
> *Woman, 52 years old*

Another participant described not knowing why exactly there is a lack of trust in the USVI system. They acknowledged that though providers on-island may have been trained in the United States, the perception is that healthcare in the United States is better; they described the importance of changing this perception to establish trust in healthcare system.

> *I guess it’s just a trust factor that even though maybe some of the doctors might come from the States and all of the nurses are probably [local university] graduates - I don’t know cause when I was in the States, you feel as if you get better care for some reason. It could be true, it could not be true but that’s how you feel and the perception - when you have a perception, you go based on the perception so if they work on changing that image, maybe you can feel more comfortable*
>
> *Woman, 51 years old*

Participants linked their identity as someone “from” the USVI, the US mainland, or other Caribbean islands to both their trust in the healthcare system and their decision-making process on where they obtained healthcare. For example, this participant sees people whom she identifies as being from USVI traveling off-island for healthcare and observing this behavior influenced her sense of trust in the healthcare system.

> *I see people leaving and going off-island for care because they themselves don’t trust the system here. I don’t belong here. Why should I trust it?*
>
> *Woman, 62 years old*

Another participant says that the difference between where her and her husband obtain healthcare is influenced by where they are from and implies the importance of “home” to her sense of trust and comfort in the healthcare system.

> *I’ve been here since [year] and I always go home. I always go off-island. I don’t have any doctors here, I only have one dentist. That’s it. All of my doctors are in the States…*
>
> *He [My husband] is a local person. The need that I feel to go up there [US mainland states] to do my health concerns, he doesn’t feel it because he is from here [USVI] and here is fine and here is everything, as a local*.
>
> *Woman, 65 years old*.

## Discussion

In the ongoing post-hurricanes rebuilding efforts, the US territories will continue to face financial and human resources challenges to its healthcare infrastructure. Our data provide USVI residents’ perspectives on why they decide to seek healthcare in other locations, mostly on the US mainland. This study builds upon previous studies that examine the factors that influence patient’s decisions on selecting where they go for healthcare and what providers they seek out.^17-22^ Such research has demonstrated the complex ways through which patients make healthcare decisions and the healthcare system factors contributing to off-island healthcare use. From this analysis, five themes emerged that illustrate healthcare system level factors that influence participants’ decisions about seeking healthcare off-island. These factors include the limited availability of services, the limit accessibility of services on-island and off-island, healthcare system interactions and experiences, healthcare system policies, and trust in the healthcare systems.

The finding that limited availability of services in the USVI results in mobility for healthcare reflects the body of literature that explores the people’s decisions to seek healthcare within other contexts. For these participants, the decisions that are influenced by limited availability are tied to their desire for choice and options, a core value for people when selecting healthcare providers in United States and other nations. The importance of choice has been described in studies exploring patient preferences in managed care settings, those with employer-based insurance, or rural locations in which people travel to more urban areas to obtain more choice.^17-20,23^ In the context of the USVI, the tension between limited availability and a desire for choice and options leads to seeking healthcare in other geographic locations. This decision is not solely based on the proximity of the closest provider but is tied to other factors, such as accessibility of healthcare in terms of cost and convenience, as well as the perception of healthcare services as having a better reputation, seen in studies related to cross-border mobility, rural-urban mobility and between private and public healthcare systems.^24-27^

Another main finding was how participants’ trust in the healthcare system influenced decisions to seek healthcare off-island.^28,29^ Some participants described how their perception of the healthcare systems was influenced in their overall distrust in institutions on-island. This finding is consistent with literature of “institutional trust,” which places the healthcare system as part of the larger society that helps create a foundation of trust for its community members.^28-30^ The healthcare system functions not only as “a guarantor of interpersonal trust but also as the foundation of trust as a property of the overall social systems.”^28-30^ Therefore, when there is a general environment of uncertainty regarding the reliability of some community institutions resulting in distrust in these institutions, this may result in an internalized perception that other settings may be inherently better.^29^ In addition, the reputation of the hospital may also influence the interpersonal relationship between the provider and patient.^31^ People may question whether the healthcare provider whom they encountered on any given day had the knowledge and skills necessary to care for their needs. Our findings also suggested that trust in the healthcare system may also stem from one’s identity as “belonging” to USVI or some other location, such that one’s identity influences how one views the healthcare system and decisions on where to obtain healthcare. Further in-depth exploration of this finding is needed.

This analysis also demonstrates the interrelation of key themes at the healthcare systems level that influence decisions to seek healthcare off-island. For example, our findings illustrate that the relationships between healthcare policies, e.g. the way insurance policies are administered on-island as compared with on the US mainland,^32,33^ influences the availability and accessibility of healthcare services and participants’ interactions with the healthcare system. Moreover, though healthcare may be available, the limited supply of services negatively affects how individuals experience the healthcare system in terms of both long wait-times for appointments and lack of choice in provider options.

These findings have implications for efforts to rebuild the healthcare system in the USVI and future research. The inter-connectedness between the themes suggests that an intervention can potentially affect multiple points that influence participants healthcare seeking decisions. For example, an investment in health information technology, such as telemedicine, as demonstrated in other isolated communities, may address some issues related to why people seek care off-islands by improving availability of providers and thereby choice.^34,35^ In addition, health information systems, e.g. an electronic health record with a patient portal, can facilitate a sense of access and coordination of care between the patient and the healthcare system, thereby improving a patient’s care experience and potentially addressing issues with cost and convenience. Revisiting healthcare workforce policies may help improve the availability and access to services, and have positive influences on people’s perspectives of and “trust” in their healthcare providers, despite known existing healthcare shortages.^36^ These offer an opportunity to rebuild trust between community and health systems by addressing trust at multiple different levels, as seen in other communities.^29^

There are several limitations to this study. As a qualitative study, our objective was not to be able to generalize results to other populations or locations. It primarily contained the perspectives of individuals who use healthcare services off-island as to understand their decisions. The perspectives of the study participants may not characterize the experiences and motivations of others on the island for seeking care on or off-island and cannot be generalized to other islands in the region. However, there may be similarities in experiences of individuals from Puerto Rico for seeking healthcare off-island, as prior to the hurricane, Puerto Rico had limited Medicaid and Medicare services as compared to the US mainland, and was facing a workforce shortage problem.^37,38^

## Conclusion

These finding elucidate the multiple health system factors that contribute to USVI residents’ decisions to seek healthcare in another geographic location, as well as illustrate the complex interactions between these factors. Future work can explore the individual and inter-personal factors, including family and social networks, that influence the decision of seeking healthcare and how this type of mobility across geographies for health services affects outcomes. As the rebuilding efforts continue in the USVI, it will be important to continue to understand the experiences of individuals seeking healthcare and their decision-making process on where to seek care. Our study indicates that these experiences are mechanisms to evaluate whether the healthcare systems are meeting the needs of patients and can provide insight into ways to improve the healthcare system.

**Figure 1.**
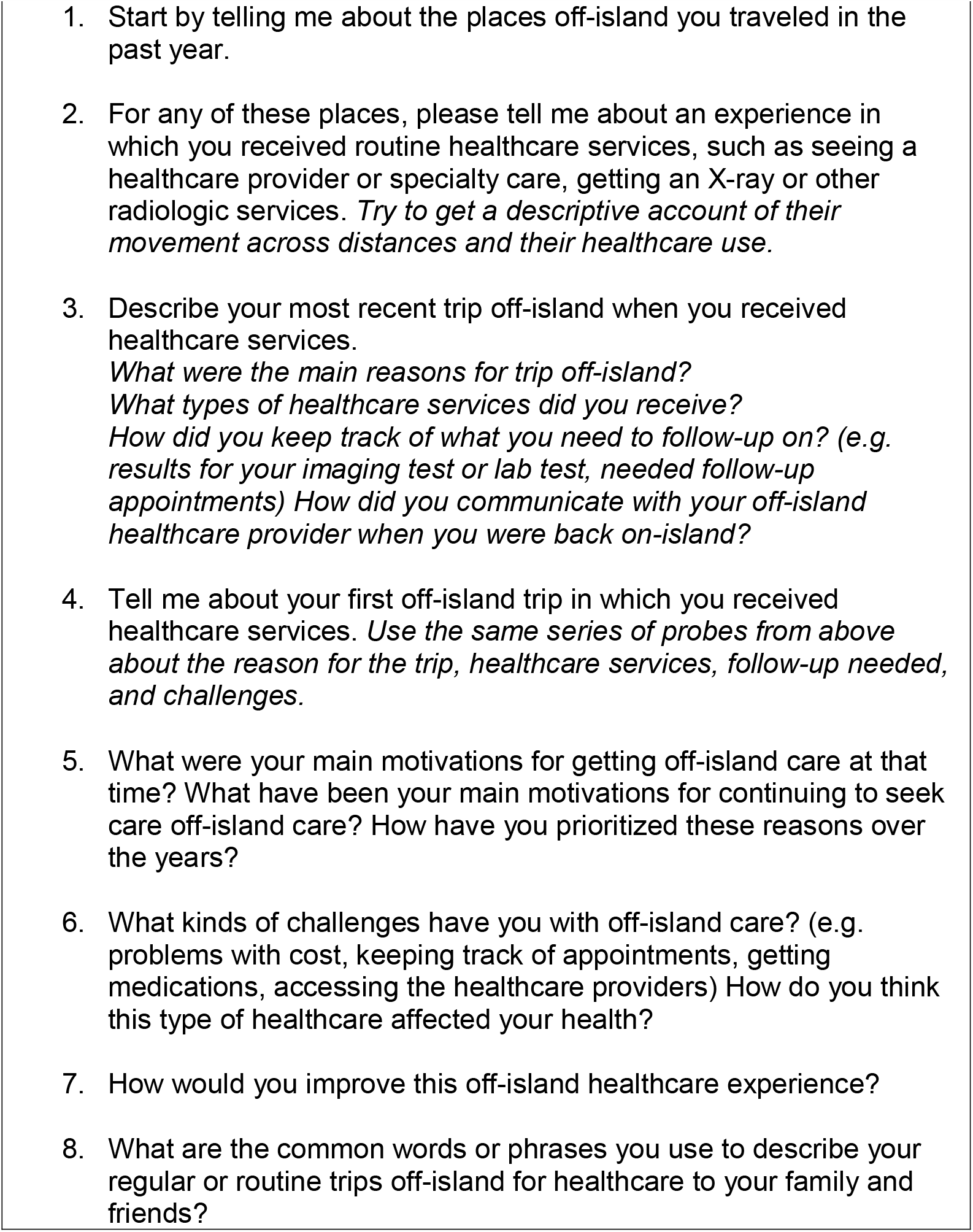
Interview Guide.

## Data Availability

Not available. Please contact the corresponding author.

